# Oral Hygiene Practices and Dental Caries Experience in Nigeria: A Systematic Review and Meta-Analysis

**DOI:** 10.64898/2026.01.30.26345076

**Authors:** Adetayo Aborisade, Amina Mohammed Ali, Chizoba Okolo, Titilola Abike Gbaja-Biamila, Folahanmi Tomiwa Akinsolu, Abideen Olurotimi Salako, Ifeoluwa Eunice Adewole, Mobolaji Timothy Olagunju, Olunike Rebecca Abodurin, George Uchenna Eleje, Ucheoma Catherine Nwaozuru, Adebola Oluyemisi Ehizele, Joanne Marie Lusher, Maha El Tantawi, Oliver Chukwujekwu Ezechi, Morẹ́nikẹ́ Oluwátóyìn Foláyan

**Affiliations:** Department of Oral Diagnostic Sciences, Bayero University, Kano, Nigeria; Department of Oral Maxillofacial Surgery, Bayero University, Kano, Nigeria; Department of Child Dental Health, Bayero University, Kano, Nigeria; Oral Health Initiative, Center for Reproductive and Population Health Studies, Nigerian Institute of Medical Research, Yaba, Lagos, Nigeria; Department of Clinical Sciences, Nigerian Institute of Medical Research, Lagos; Department of Public Health, Faculty of Basic Medical and Health Sciences, Lead City University, Ibadan; Department of Epidemiology and Health Statistics, Nanjing Medical University, China; Lagos State Health Management Agency, Lagos; Department of Obstetrics and Gynaecology, Nnamdi Azikiwe University Teaching Hospital, Nnewi, Nigeria; Effective Care Research Unit, Department of Obstetrics and Gynaecology, Nnamdi Azikiwe University, Awka, Nigeria; Department of Community Medicine and Primary Healthcare, Nnamdi Azikiwe University Teaching Hospital, Nnewi, Nigeria; Wake Forest School of Medicine, North Carolina, USA; Department of Periodontics, School of Dentistry, College of Medical Sciences, University of Benin, Benin City, Nigeria; Provost’s Group, Regent’s University London, UK; Department of Pediatric Dentistry and Dental Public Health. Faculty of Dentistry, Alexandria University, Egypt; Department of Child Dental Health, Obafemi Awolowo University, Ile-Ife, Nigeria

**Keywords:** Oral hygiene practices, dental caries, tooth brushing, Nigeria, meta-analysis

## Abstract

**Background:** Oral hygiene is linked with dental caries experience. This systematic review and meta-analysis assessed the associations between oral hygiene status, the frequency of tooth brushing, and the prevalence of dental caries in Nigeria. Tools used for maintaining oral hygiene were also identified.

**Methods:** Registered with PROSPERO (CRD42022367763), this review searched PubMed, Web of Science, Scopus, African Journals Online, African Index Medicus, and Google Scholar in January 2026. Observational studies and clinical trials reporting baseline caries prevalence were included. There was no language restriction. Studies were excluded if they did not provide information on the sample size, had no study outcome data, or featured duplicate samples, and if they were review articles, systematic reviews and meta-analyses, case reports, case series, in vitro studies, commentaries/letters (editorials, opinion pieces) devoid of primary data. Pooled odds ratios (ORs) were estimated using random-effects models. Subgroup analyses were conducted by dentition type, geopolitical zone, study design, publication year, mean age, and sample size.

**Results:** Twenty-three cross-sectional studies were included, of which 20 (86.9%) were conducted in Southern Nigeria. After removing an influential outlier, poor oral hygiene was associated with a 38% reduction in caries odds (OR 0.62, 95% CI 0.46–0.84). Brushing at least twice daily was strongly associated with reduced caries (OR 0.01, 95% CI 0.00–0.01). No significant association was found between the type of cleaning device and caries prevalence. Subgroup analyses identified dentition type and publication year as significant moderators. Heterogeneity ranged from low to moderate, and no publication bias was detected for primary associations. The most common cleaning tool was a toothbrush with toothpaste, though chewing sticks, cotton wool, and other traditional tools were also reported.

**Conclusion:** Twice-daily tooth brushing is strongly associated with lower caries prevalence in Nigeria. Poor oral hygiene was linked to reduced caries odds in pooled analysis, a finding that may reflect socio-economic and dietary confounding. The type of cleaning tool was not significantly associated with caries risk, highlighting the importance of brushing frequency and technique over tool type. Future research should prioritize Northern Nigeria to address the geographic skewness of the data and improve national representativeness.

## Introduction

Oral hygiene is essential for oral and general health. It encompasses a range of preventative practices that individuals adopt to maintain the cleanliness and overall health of the oral cavity. [1–3]. Key oral hygiene methods include regular tooth brushing with a toothbrush and toothpaste, which removes dental plaque and food particles, prevents periodontal diseases, and dental caries. [4]. Other practices, such as dental floss or interdental cleaning, antimicrobial rinses, and tongue cleaning, complement tooth brushing in controlling plaque accumulation and maintaining oral health. [5, 6]. Furthermore, oral hygiene includes thorough cleaning of the dorsum of the tongue using a tongue scraper or a toothbrush, which helps to eliminate bacteria and debris from the oral cavity because the tongue can be a reservoir for oral bacteria [7–9].

The link between oral hygiene and dental caries is well-established, as dental plaque plays a central role in initiating and advancing dental caries lesions [10, 11], which suggests that efforts to control plaque would positively affect the prevention of dental caries [12]. Regular toothbrushing, fluoridated toothpaste, and dental floss help remove plaque and reduce the availability of fermentable carbohydrates for bacterial acid production. [6, 13–15]. Oral hygiene practices can reduce the risk of developing dental caries when combined with professional dental care. [11, 14, 16, 17]. Proper oral hygiene removes plaque from tooth surfaces. [10, 18], which prevents the initiation of enamel demineralization caused by acid produced by bacterial fermentation of carbohydrates in the oral cavity [3, 19].

Globally, untreated dental caries in permanent teeth remains the most prevalent health condition, affecting an estimated two billion people in 2022 [20]. Despite this burden, there is limited context-specific evidence from Nigeria. The most recent national survey on dental caries in Nigeria was conducted in 1995, and a national survey on oral hygiene practices was last conducted in 2014 [21] Since then, major socio-economic and demographic shifts, urbanization, dietary changes (including increased sugar consumption), and disparities in access to preventive oral health services have likely altered the relationship between oral hygiene practices and dental caries. Furthermore, regional diversity in cultural practices, such as the use of chewing sticks alongside toothbrushes, necessitates evidence that reflects both modern and traditional practices. Without updated, nationally representative evidence, policymakers and oral health professionals lack the data needed to design effective, culturally relevant, and equitable interventions for dental caries prevention.

In the absence of national surveillance, a systematic review can address the critical evidence gap and offer insights to guide oral health policies, prevention strategies, and public health programs in line with the United Nations Sustainable Development Goal 3 on good health and well-being [22]. This systematic review and meta-analysis therefore aimed to synthesize available evidence on the association between oral hygiene practices and dental caries in Nigeria and to provide a national estimate from published observational studies.

## Methods

### Study protocol

This systematic review and meta-analysis were initially registered with PROSPERO (CRD42022367763) in 2022 and updated on the 24^th^ of January 2026. The study was reported in accordance with the Preferred Reporting Items for Systematic Reviews and Meta-analyses (PRISMA) statement and checklist. [23]. Two blinded reviewers performed each review stage, and disagreements were resolved through discussions with a third reviewer.

### Research questions

The following research questions guided the study: 1) Is there an association between oral hygiene status and dental caries? 2) Is there an association between the frequency of tooth brushing and the prevalence of dental caries? 3) What tools were used by participants for maintaining oral hygiene?

### Search strategy

Five databases (PubMed, Web of Science, Scopus, African Journals Online, and African Index Medicus) and Google Scholar were searched for relevant articles published with no language restriction. The initial search syntax was developed for PubMed and later adapted to fulfill the unique search criteria of the other databases (Supplemental File 1).

To ensure a comprehensive search, grey literature was sought from the libraries of the National Postgraduate Medical College of Nigeria and the West African College of Surgeons. In addition, the reference lists of all retrieved articles, systematic reviews, and meta-analyses were examined. To identify further sources, study investigators were contacted directly for inaccessible publications and for clarification on missing data, theses, and unpublished manuscripts.

### Inclusion and Exclusion Criteria

All published and unpublished studies, including hand searches conducted in Nigeria between January 2001 and December 2025, that reported on associations between oral hygiene and the prevalence of dental caries were eligible for study inclusion. Study designs eligible for inclusion were cross-sectional, cohort, and case-control studies. Clinical trials that provided relevant baseline prevalence data were also included. Studies were also included if they presented available data for at least one of the primary outcomes indicated in Table 1, whose reported odds ratio (OR) and 95% confidence interval (CI) were analyzed by univariate or multivariate analyses, and studies whose data could be analyzed to generate variables for the current study.

**Table 1:** Eligibility Criteria using PECOS (Population, Exposure, Comparisons, Outcomes, Studies) framework.

|  |  |
| --- | --- |
| Population | Individuals residing in Nigeria |
| Exposure | Oral hygiene practices (tooth brushing frequency and tooth cleaning methods - toothbrush, chewing stick, or other devices), and oral hygiene status. |
| Comparators | Variations in oral hygiene practices (e.g., brushing $\geq 2$ times/day vs. once/day; good vs. poor hygiene; toothbrush vs. chewing stick). |
| Outcomes | Presence of dental caries. |
| Studies | Studies reporting prevalence data. |

Studies were excluded if they did not provide information on the sample size, had unavailable outcome data, or featured duplicate samples. Review articles, systematic reviews and meta-analyses, case reports or case series, in vitro studies, commentaries/letters (editorials, opinion pieces), devoid of primary data, were excluded. Studies with overlapping data from other included studies were also excluded. Excluded studies are reported in Supplemental File 2.

### Selection of studies

Studies were screened using the PECOS framework (Population, Exposure Comparators, Outcomes, Time, Studies). Table 1 presents the PECOS framework used for this study. [24]. In this review, dental caries was the primary outcome of interest. Oral hygiene practices were considered the exposures, and different categories of practices served as comparators. This distinction was made explicit to ensure clarity in the analytic framework and to avoid misinterpretation of dental caries as an exposure or risk factor.

Three authors (AA, AMA, and CO) independently reviewed the titles and abstracts of each study that met the inclusion criteria after removing duplicates and downloading them to the reference management software EndNote 7.8. Studies that did not meet the inclusion criteria and those where the full text was unavailable were excluded. Two reviewers (AA and AMA) independently assessed the eligibility of the retrieved manuscripts, and any disagreements were resolved by discussion or recourse to a third reviewer (MOF).

### Data Extraction

Four independent reviewers (AA, TAG, IEA, and ORA) used a pretested data extraction form prepared in Microsoft Excel to independently extract information related to the author’s name and year of publication. In addition, specific details about the study design, location, and study setting were captured. Details about the study participants – sample size, age distribution, sex of participants, and other unique characteristics – were extracted. Information about participants’ oral hygiene practices was also extracted. These details included measures of oral hygiene status, frequency and duration of hygiene practices, and the tools or methods employed for oral hygiene. Simultaneously, dental caries assessment data were cataloged, highlighting each study’s tools or methods for assessing dental caries. The percentage of participants with dental caries was also extracted. Lastly, the results or association measures, whether in the form of prevalence or statistical ratios indicating the associations between oral hygiene practices and dental caries, were recorded. Any discrepancies were resolved by a fifth reviewer (MOF).

### Quality and Risk of Bias Assessment

Four independent reviewers (AA, TAG, IEA, and ORA) assessed the methodological quality and risk of bias in the included studies, with discrepancies resolved by a fifth reviewer (FTA) using an adapted version of the risk of bias tool for prevalence studies according to the modified Joana Briggs Institute Assessment for Risk of Bias [25]. This risk of bias tool, designed for prevalence studies, was applied as the most appropriate quality assessment approach because the Odds Ratios (ORs) synthesized in the meta-analysis were calculated directly from raw prevalence counts (number of events and non-events) reported in the included cross-sectional studies. The validity of these ORs is fundamentally dependent on the methodological quality of the prevalence data. The total score ranged from 0 to 9, with the overall score categorized as follows: 0–3: “high risk,” 4–6: “moderate risk,” and 7–9: “low risk” of bias.

### Certainty of evidence

The certainty of evidence for each outcome using the Grading of Recommendations Assessment, Development, and Evaluation (GRADE) approach was not conducted because, under this framework, cross-sectional studies are rated as low certainty evidence [26, 27]. The data included in this study were all cross-sectional in design.

### Sensitivity analysis

A sensitivity analysis was planned to be performed with leave-one-out analysis to investigate the impact of risk of bias on the test of associations and to uncover a potential source of heterogeneity.

### Assessment for publication bias

Symmetry of the funnel plot was planned to assess the publication bias of the articles included in the systematic review. Egger’s regression test was also utilized to assess publication bias objectively with significance at p<0.05.

### Statistical Analysis

All analyses were conducted using R and RStudio Version 4.5.1 (“Great Square Root”). A descriptive analysis was first undertaken to show the distribution of studies by year of publication, study design, geopolitical region, number of male and female participants, and type of dentition studied: primary (0 –5 years), mixed (6–11 years), and permanent (≥12 years). We also described the number of studies reporting an association between oral hygiene practices and dental caries prevalence by geographic region and dentition type.

This meta-analysis applied strict inclusion criteria, selecting only studies that reported definitive, binary data on the presence or absence of dental caries. This ensured an objective and direct assessment of the relationship between oral hygiene practices and caries, aligning with standard epidemiological methods for evaluating disease prevalence and risk factors. Pooled estimates were generated using the Mantel-Haenszel method. Heterogeneity was evaluated using Cochran’s Q test, the I² statistic, and inspection of forest plots. The independent variables were oral hygiene practices (tooth cleaning method, brushing frequency, and oral hygiene status), while the dependent variable was dental caries prevalence. Odds ratios (ORs) with 95% confidence intervals (CIs) were used as the summary measure, categorizing outcomes into dental caries present versus absent. These ORs were calculated from extracted frequencies and denominators reported in the studies, rather than adopting published effect sizes. Because the synthesis was based on prevalence data, a risk of bias tool designed for prevalence studies was applied as the most appropriate quality assessment approach. A p-value <0.05 for the Q test was considered statistically significant, and I² values above 50% indicated substantial heterogeneity. Negative I² values were treated as zero [28, 29], following Cochrane Handbook guidance [30, 31]. Studies with no events in either arm were excluded from the meta-analysis. [29–32].

Associations between oral hygiene practices and dental caries prevalence were evaluated using moderator analysis, including subgroup analysis and univariate meta-regression. When study heterogeneity is high, subgroup analyses were conducted based on the type of dentition (primary, mixed, permanent), geopolitical zone (South and Northern), and study design (population-based, school-based, or other). Univariate meta-regression examined the year of publication, study sample size, and participants’ mean age in each study. Sensitivity analyses involved excluding studies with a high risk of bias and sequentially removing individual studies to assess their influence on pooled estimates. Publication bias was assessed using funnel plots and Egger’s test when at least ten studies reported the same outcome [33, 34].

## Results

### Selection of studies

As shown in Figure 1, 1422 records were retrieved. After removing 414 duplicates, 1008 records remained for eligibility screening. Of these, 935 studies were excluded based on their titles and abstracts. After reviewing the full-text records, 23 met the inclusion criteria [5, 8, 15, 18, 35–53].

**Figure 1:**
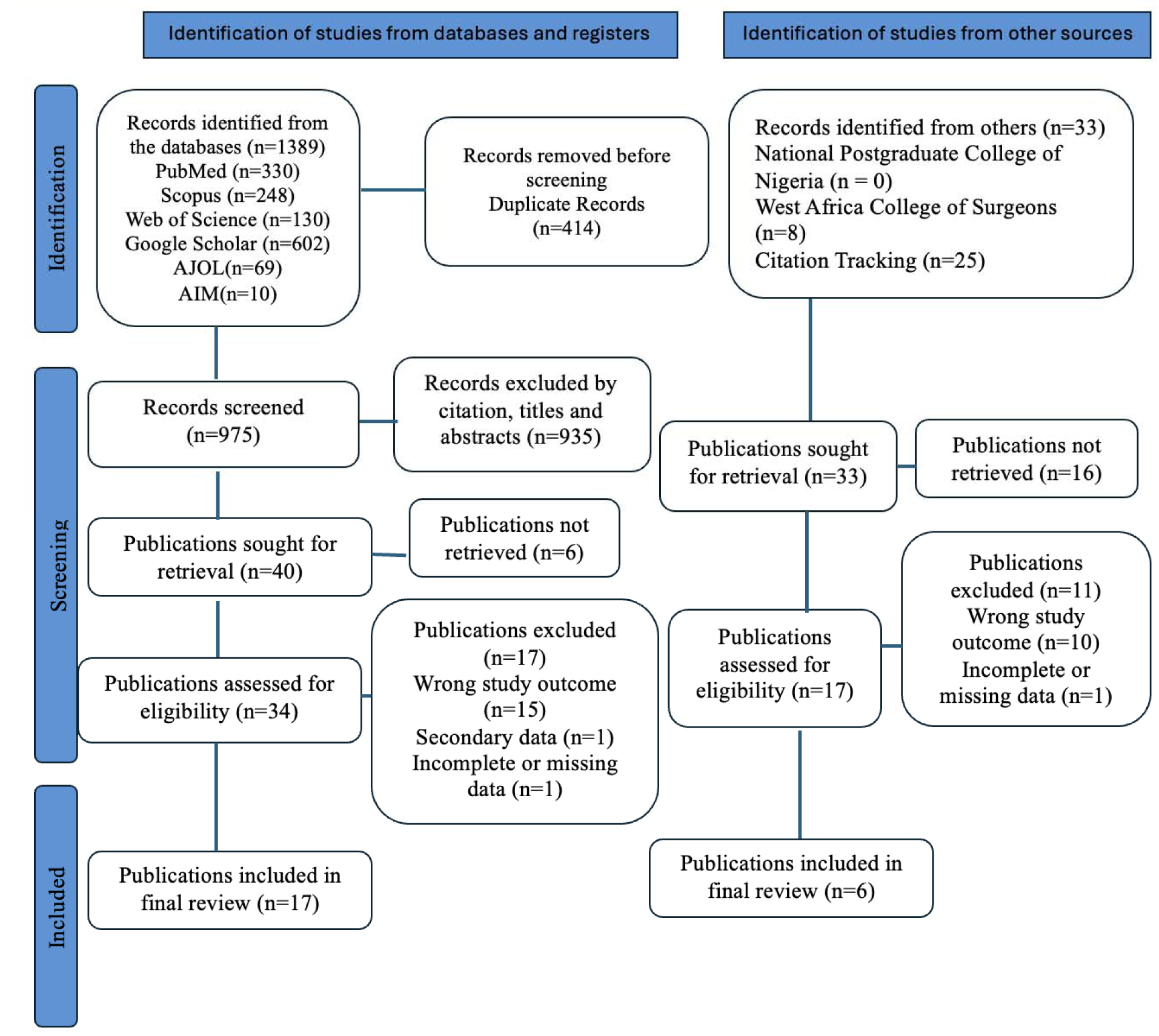
PRISMA flow diagram for the search results

### Characteristics of included studies

The characteristics of the 23 included studies are detailed in Table 2. The participant population size in each study ranged from 124 to 2,107 individuals, culminating in a combined total of 15,797 participants. The aggregated sample comprised 7838 males and 7959 females with participants’ ages ranging from six months to 64 years.

**Table 2:** Characteristics of Included Studies.

| S/No | Study Reference | Title | Study Design | City/State | Region | Sample Size | Age Range | Gender Distribution (M/F) |
| --- | --- | --- | --- | --- | --- | --- | --- | --- |
| 1 | Sowole et al., 2007 [15] | Dental Caries Pattern and Predisposing Oral Hygiene Related Factors in Nigerian Preschool Children | Cross-sectional | Lagos | Southwest | 389 | 6-71 months | 217 / 172 |
| 2 | Okoye et al., 2011 [35] | Prevalence of Dental Caries in a Nigerian Rural Community: A Preliminary Local Survey | Cross-sectional | Enugu | Southeast | 301 | 11-16 years | 100 / 201 |
| 3 | Ajayi and Abidemi-Solanke, 2014 [36] | Socio-behavioural risk factors of dental caries among selected adolescents in Ibadan, Nigeria | Cross-sectional | Ibadan | Southwest | 914 | 10-19 years | 451 / 463 |
| 4 | Folayan et al., 2015 [37] | Prevalence And Early Childhood Caries Risk Indicators in Preschool Children in Suburban Nigeria | Cross-sectional | Osun | Southwest | 497 | 6-71 months | 266 / 231 |
| 5 | Olatosi et al., 2015 [38] | The Prevalence of Early Childhood Caries and Its Associated Risk Factors Among Preschool Children Referred to a Tertiary Institution | Cross-sectional | Lagos | Southwest | 202 | 6-71 months | 44 / 158 |
| 6 | Olabisi et al., 2015 [8] | Prevalence Of Dental Caries and Oral Hygiene Status Of A Screened Population in Port Harcourt, Rivers State, Nigeria | Cross-sectional | Rivers | South-South | 288 | 15-64 years | 133 / 155 |
| 7 | Onyejaka and Amobi, 2016 [39] | Risk Factors of Early Childhood Caries among Children in Enugu, Nigeria | Cross-sectional | Enugu | Southeast | 429 | 9 months - 5 years | 226 / 203 |
| 8 | Akinyamoju et al., 2018 [18] | Dental Caries and Oral Hygiene Status: Survey of Schoolchildren in Rural Communities, Southwest Nigeria | Cross-sectional | Ogun | Southwest | 778 | 7-17 years | 424 / 354 |
| 9 | Oyedele et al., 2018 [40] | Impact of Oral Hygiene and Sociodemographic Factors on Dental Caries in a Suburban Population in Nigeria | Cross-sectional | Osun | Southwest | 2,107 | 8-16 years | 982 / 1125 |
| 10 | Kolawole et al., 2019 [5] | Association Between Malocclusion, Caries, And Oral Hygiene in Children 6 To 12 Years Old Residents in Suburban Nigeria | Cross-sectional | Osun | Southwest | 495 | 6-12 years | 242 / 253 |
| 11 | Abiola et al., 2019 [41] | Dental Caries Occurrence and Associated Oral Hygiene Practices Among Rural and Urban Nigerian Preschool Children | Cross-sectional | Lagos | Southwest | 404 | 18-60 months | 208 / 196 |
| 12 | Nwathor et al., 2019 [42] | Oral Hygiene Status, Practices, and Awareness of Medium Security Prison Inmates in Northeastern Nigeria | Cross-sectional | Bauchi | Northeast | 280 | <30->50 years | 276 / 4 |
| 13 | Disa et al., 2019 [43] | Predictors of dental caries among adults and adolescents in a dental clinic in North-eastern Nigeria | Case-control | Yobe | Northeast | 124 | 15-63 years | 59 / 65 |
| 14 | Oyapero et al., 2020 [44] | Association Between Dental Caries, Odontogenic Infections, Oral Hygiene Status, and Anthropometric Measurements of Children in Lagos, Nigeria | Cross-sectional | Lagos | Southwest | 278 | 1-15 years | 132 / 146 |
| 15 | Arowolo, 2020 [45] | Determination Of the Association Between Nutritional Status and Dental Caries In 6-16-Year-Old School Children In Ile – Ife | Cross-sectional | Osun | Southwest | 1,502 | 6-16 years | 689 / 813 |
| 16 | Olatosi et al., 2020 [46] | Disparities in Caries Experience and Socio-Behavioural Risk Indicators Among Private School Children in Lagos, Nigeria | Cross-sectional | Lagos | Southwest | 592 | 5-16 years | 307 / 285 |
| 17 | Folayan et al., 2021 [47] | Associations Between a History of Sexual Abuse and Dental Anxiety, Caries Experience, and Oral Hygiene Status Among Adolescents in Sub-Urban South West Nigeria | Cross-sectional | Osun | Southwest | 1,056 | 10-19 years | 598 / 458 |
| 18 | Onyejaka et al., 2021 [48] | Prevalence and Associated Factors of Dental Caries Among Primary School Children in Southeast Nigeria | Cross-sectional | Enugu | Southeast | 657 | 5-17 years | 316 / 341 |
| 19 | Idowu et al., 2021 [49] | Dental Caries Prevalence, Restorative Needs and Oral Hygiene Status in Adult Population: A Cross-sectional Study among Nurses in Jos University Teaching Hospital, Jos, Nigeria | Cross-sectional | Jos | Northcentral | 251 | 9–64 years | 77 / 174 |
| 20 | Oyedele et al., 2021 [50] | Comparison of Dental Caries and Oral Hygiene Status of Children in Suburban areas with those in the Rural Population of Southwestern Nigeria | Cross-sectional | Osun/Ogun | Southwest | 1,397 | 8-12 years | 706 / 691 |
| 21 | Folayan et al., 2021 [51] | Individual and familial factors associated with caries and gingivitis among adolescents resident in a semi-urban community in South Western Nigeria | Cross-sectional | Osun | Southwest | 1,472 | 10-19 years | 846 / 626 |
| 22 | Oyedele et al., 2022 [52] | Associations Between Dental Caries, Oral Hygiene Status, And Oral Health Practices of First-Year Undergraduates in A Private University in Nigeria | Cross-sectional | Ogun | Southwest | 1,164 | 15-23 years | 443 / 721 |
| 23 | Afolabi, 2023 [53] | Comparative assessment of the oral health status of children aged 4-16 years with and without sickle cell anaemia resident in Ile-Ife and Ilesha, Osun State, Nigeria | Case- control | Osun | Southwest | 220 | 6-16 years | 96 / 124 |

The publication timeline of the included studies spans 16 years, from 2007 to 2023, and can be categorized into distinct periods. (1) Early Period (2007-2015): Six studies (26.1%) were published during this foundational phase [8, 15, 35, 36, 37, 38]. (2) Middle Period (2016-2020): Ten studies (43.5%) were published, indicating an increase in research activity [5, 18, 39, 40, 41, 42, 43, 44, 45, 46]. (3) Recent Period (2021-2023): Seven studies (30.4%) were published in the most recent years, demonstrating sustained interest in the topic [47, 48, 49, 50, 51, 52, 53]. All 23 studies employed an observational design. Twenty-one studies (91.3%) were cross-sectional surveys [5, 8, 15, 18, 32–42, 44–52], while two studies (8.7%) utilized a case-control design [43, 53].

Geographically, the studies showed a significant concentration in Southern Nigeria. Twenty studies (86.9%) were conducted in the southern region, compared to three studies (13.1%) from Northern Nigeria. Within Southern Nigeria, the Southwest zone was the most researched, with 16 studies (69.6% of the total). These were conducted across Lagos [15, 38, 41, 44, 46], Osun [5, 37, 40, 45, 47, 50, 51, 53], Ogun [18, 50], 52], and Oyo (Ibadan) [36] states. Three studies (13.0%) were conducted in the Southeast (Enugu) [35, 39, 48], and one study (4.3%) was from the South-South (Rivers) region [8]. In Northern Nigeria, two studies were from the Northeast (Bauchi and Yobe) [42, 43], and one study was from the Northcentral (Jos) zone [49].

### Oral hygiene status, frequency, and devices used for oral hygiene

#### Oral Hygiene Status Assessment

Oral hygiene status was assessed in 14 (60.9%) of the 23 studies for association with the prevalence of dental caries. The Simplified Oral Hygiene Index (OHI-S) was the predominant tool, used in 12 (85.7%) of the studies [5, 15, 36, 37, 39, 40, 41, 48, 49, 50, 52, 53], and one study used the Plaque Index [51]. Most studies that used the OHI-S reported the distribution of participants across categories of good, fair, and poor oral hygiene. One study did not specify its tool for OHI measurement [40], while another presented mean OHI-S scores stratified by urban/rural setting, gender, and age [41].

#### Measures of Oral Hygiene Practices

Studies assessed various dimensions of oral hygiene behaviour:

1. **Frequency of Mouth Cleaning:** Reported in 17 studies [15, 18, 35–39, 41–43, 45–47, 49, 51–53]. The reported frequencies ranged from ‘none’ [15, 39], rarely [42], and ‘occasionally’ [18, 35] to once daily, twice daily, and more than twice daily [15, 18, 34, 35, 36, 37, 38, 40, 41, 42, 44, 45, 46, 48, 50, 51, 52].
2. **Devices Used for Mouth Cleaning:** Documented in 14 studies [15, 18, 35, 36, 38, 41, 42, 43, 45, 47, 49, 51, 52, 53]. A wide array of devices was reported, including toothbrushes with toothpaste (most common), chewing sticks, cotton wool, dental floss, gauze, fingers, herbs, and traditional agents like glycerine or ground glass.
3. **Other Behavioural Variables:** Several studies explored additional factors, including the individual responsible for cleaning (child or caregiver) [38], supervision of brushing [38, 41], age at onset of oral cleaning [38], timing of cleaning (e.g., before/after meals) [42, 49, 52], and the frequency of changing cleaning devices [18, 41, 43].

#### Dental Caries Assessment

All 23 studies used the DMFT/dmft indices to assess caries prevalence and severity. Two studies additionally employed the PUFA/pufa index to measure the clinical consequences of untreated caries [5, 43]. The assessed dentitions were: primary dentition (7 studies) [15, 37, 38, 39, 41, 45, 53], permanent dentition (9 studies) [8, 35, 42, 43, 47, 49, 51, 52, 53], and mixed dentition (8 studies) [5, 18, 36, 40, 44, 46, 48, 50]. The reported caries prevalence ranged widely from 2.2% to 79.1%.

#### Association between Oral Hygiene Status and Dental Caries

Of the 19 studies that assessed oral hygiene status, 14 (73.7%) investigated its association with caries [5, 15, 36, 37, 39, 40, 41, 44, 48, 49, 50, 51, 52, 53]. A consistent finding was that poor oral hygiene status was significantly associated with a higher prevalence or severity of caries in eight of these studies [5, 15, 39, 40, 44, 45, 52, 53]. For example, poor oral hygiene increased the odds of caries by factors ranging from 1.8 to 15.5 [5, 45, 52]. Conversely, good [48] and fair [39, 40, 48] oral hygiene were associated with a lower risk for caries in several studies. Fair oral hygiene was also associated with increased odds of caries [40, 52]. Some studies reported a dose-response relationship, where the risk of caries increased with higher (worse) OHI-S scores [5, 41]. Four studies found no significant association [36, 49, 51, 53].

#### Association between Oral Hygiene Practices and Dental Caries

The evidence for associations between specific oral hygiene behaviours and caries was less consistent:

- **Frequency of Cleaning:** Most studies (8/10) found no significant association between brushing frequency (e.g., once vs. twice daily) and caries prevalence [18, 35, 36, 42, 46, 47, 51, 53]. Two studies reported protective effects for brushing at least twice daily [43, 45].
- **Cleaning Devices:** The use of a toothbrush with fluoride toothpaste was generally associated with lower caries experience compared to traditional methods like using glycerine, which was linked to significantly higher odds of caries (OR: 17.7) [38]. One study found that chewing stick users had more caries than toothbrush users among males [35]. However, most studies found no significant association for the use of toothbrushes, chewing sticks, floss, or other specific devices when analyzed independently [18, 36, 42, 51].
- **Other behavioural Variables:** Factors such as the supervisor of brushing, age of onset of cleaning, timing of cleaning [42], and frequency of changing devices [18] were not associated with caries prevalence. One study found that not cleaning after every meal was protective [43].

#### Risk of Bias Assessment of Included Studies

All but one of the included studies [43] were determined to be of low risk of bias, scoring 7–9 on the quality scale [5, 8, 15, 18, 35–42, 44–53] (See Supplemental File 3)

**Table 3:**
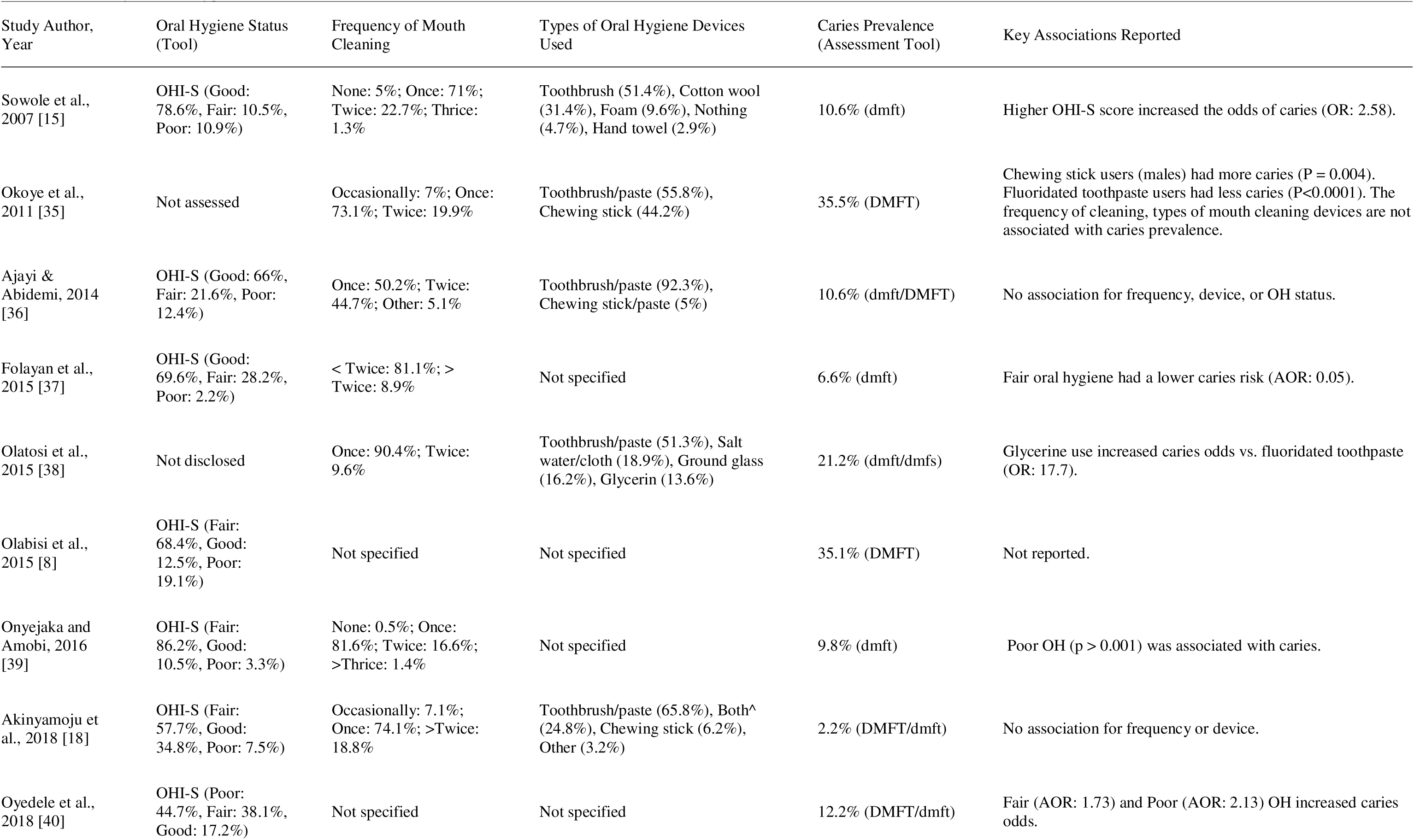
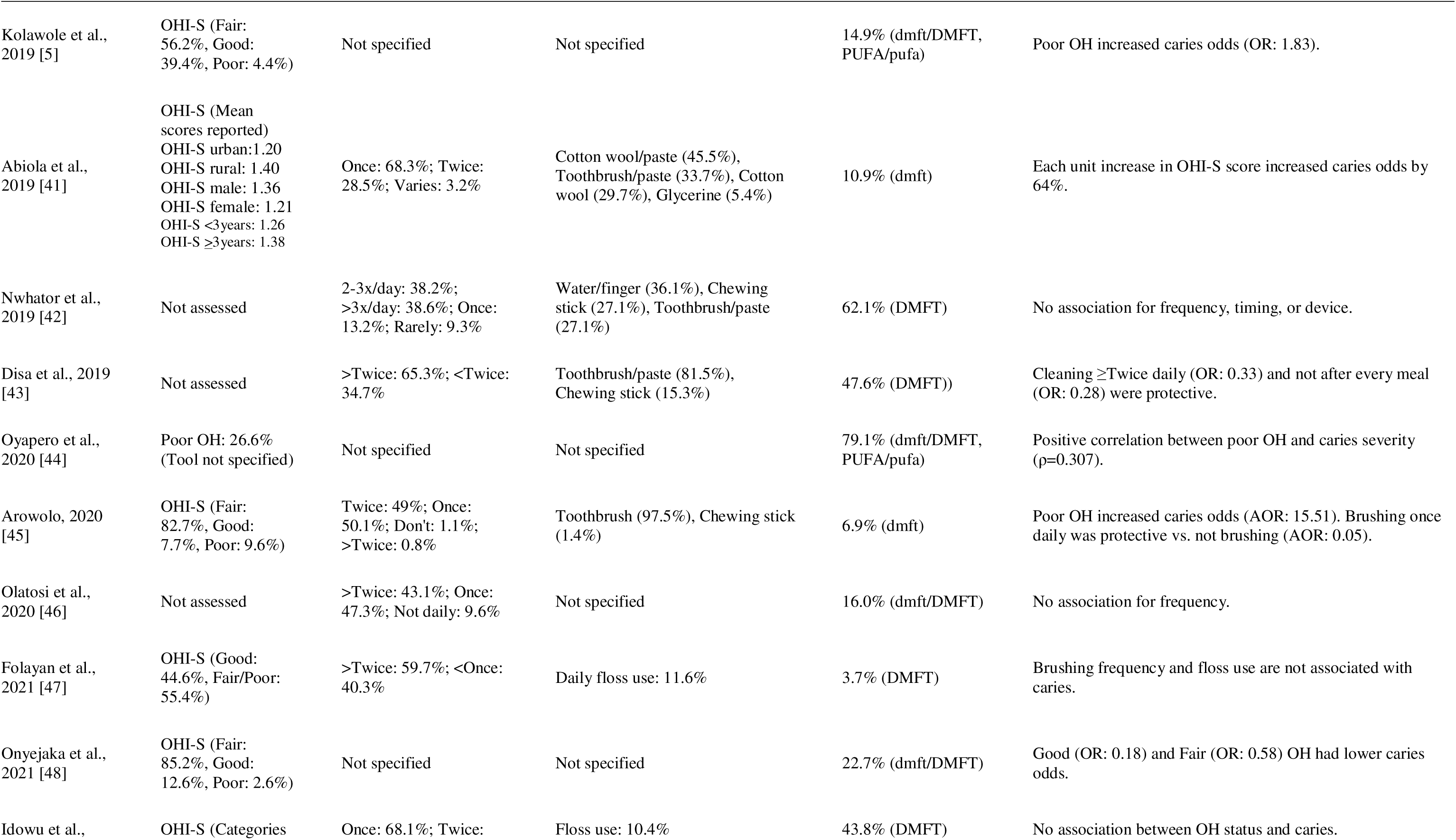
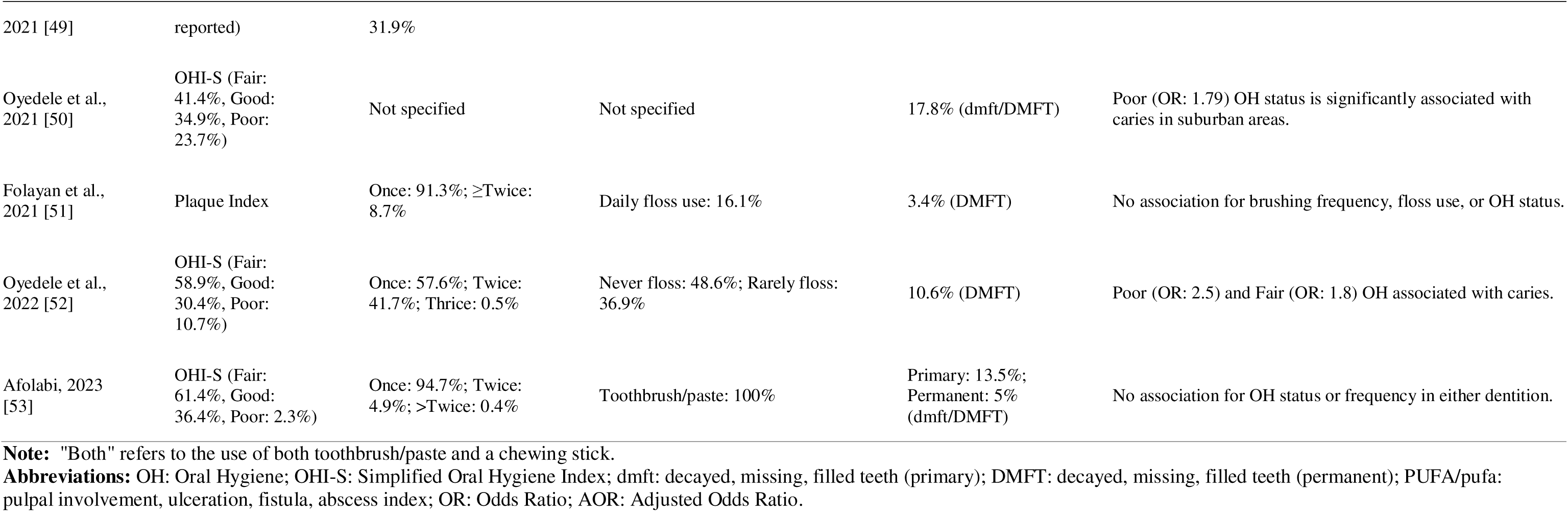
Summary of Oral Hygiene Practices and Associations with Dental Caries in Included Studies.

#### Association between oral hygiene status and dental caries

Pooled data from 10 studies initially suggested poor oral hygiene was associated with lower caries odds (OR 0.52, 95% CI: 0.32–0.85; p=0.01). After removing a study that was an influential outlier [47], the analysis showed a significant 38% decrease in caries prevalence associated with poor oral hygiene (OR 0.62, 95% CI 0.46–0.84; p = 0.006), with moderate heterogeneity (I² = 44.6%).

**Figure 2:**
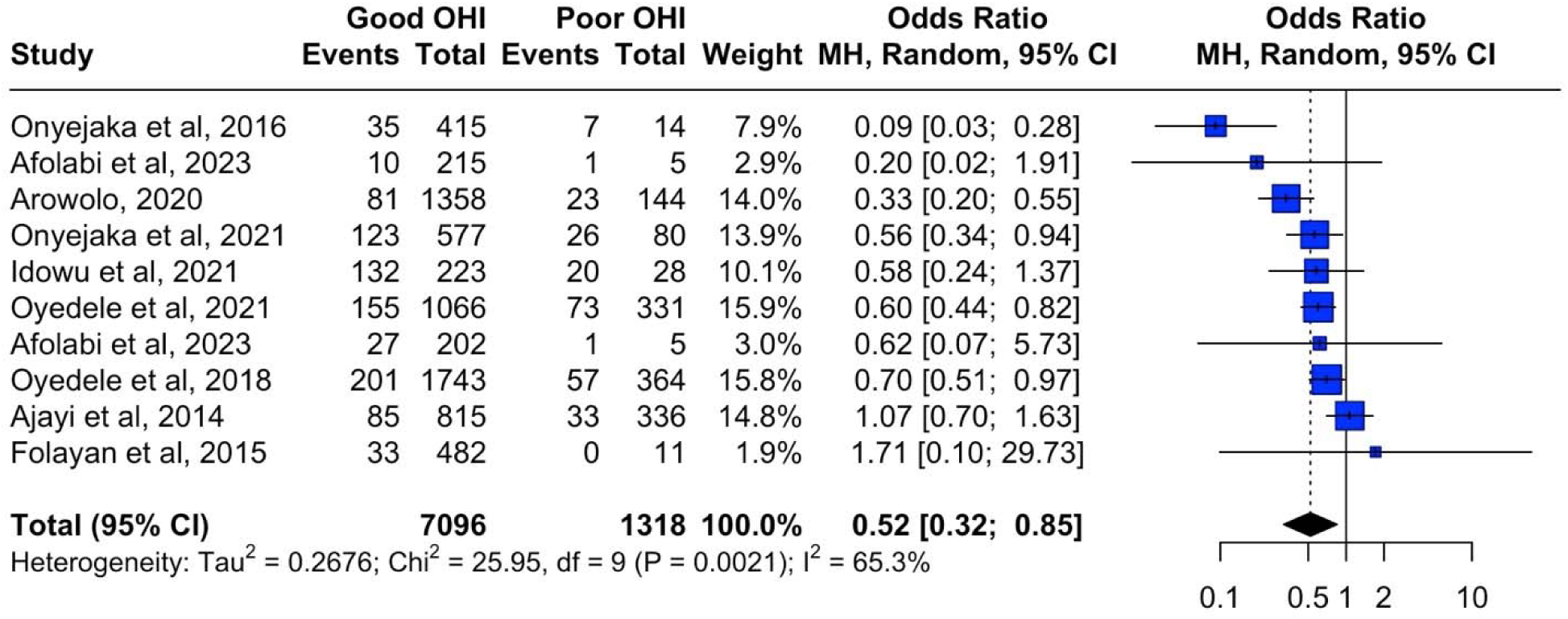
Forest plot showing the meta-analysis of oral hygiene status and caries prevalence

**Figure 3:**
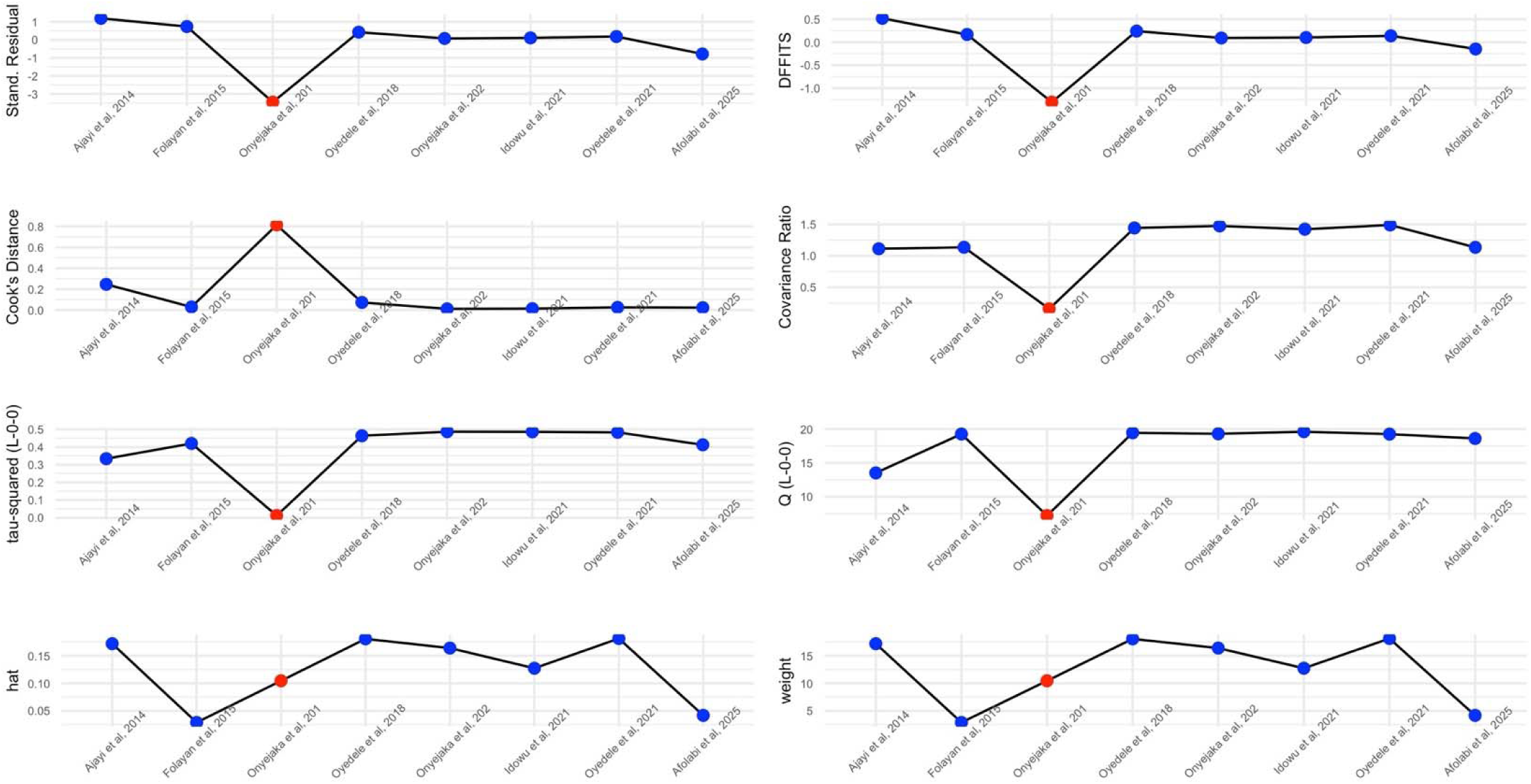
Influential plot showing Onyejaka et al. [47] was an influential study

#### Subgroup analyses

Subgroup analysis using the type of dentition showed that the type of dentition was a significant moderator in the association of caries prevalence with oral hygiene status (chi^2^=7.46, p=0.02). Oral hygiene status was associated with a 64% reduction in odds for caries in the primary dentition and a 49% reduction in caries prevalence in the permanent dentition (Figure 4). Odds reduction observed with good oral hygiene in the mixed dentition was 39% for caries prevalence.

**Figure 4:**
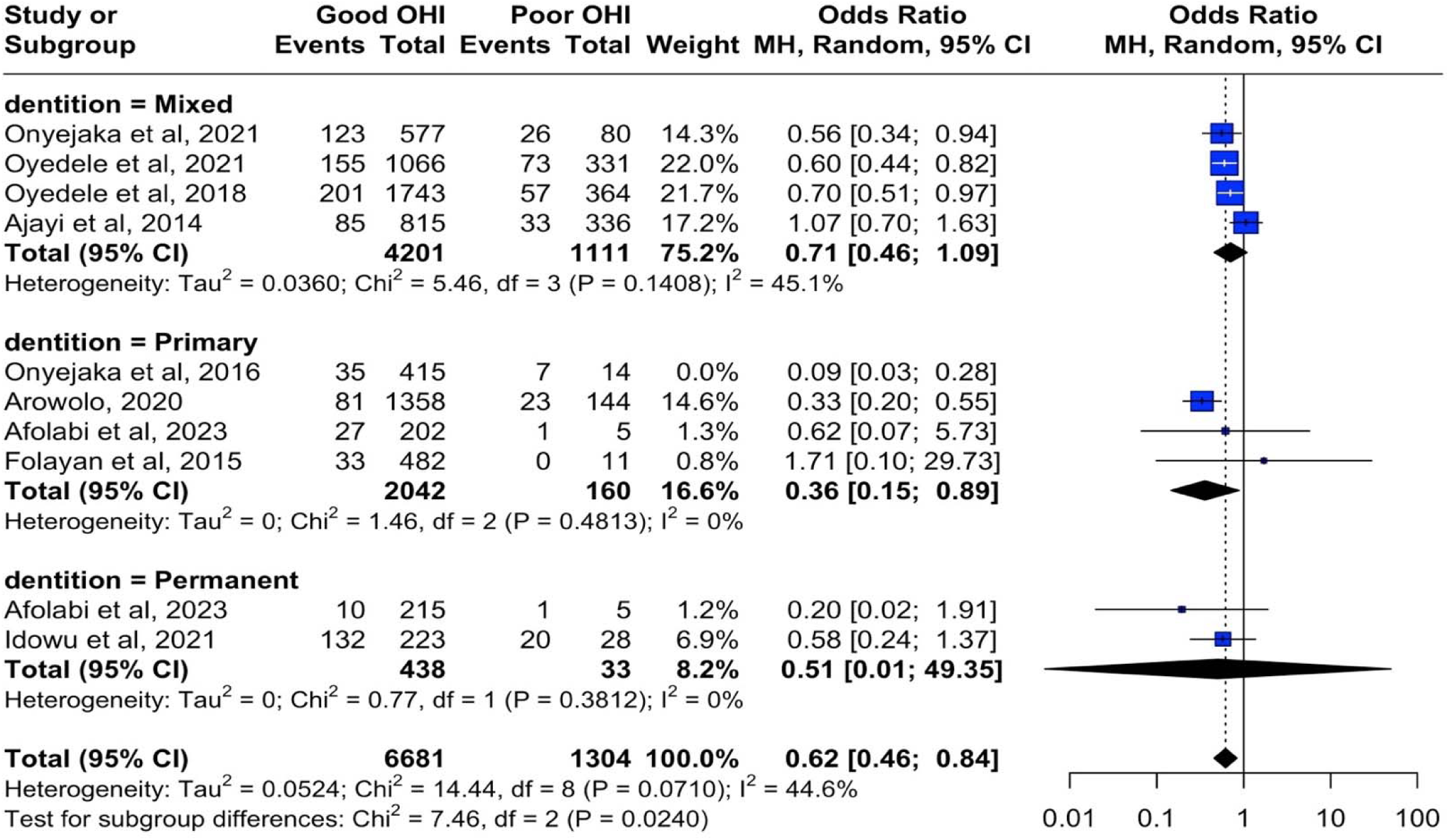
Forest plot showing subgroup analysis by type of dentition for pooled prevalence and OHI

Subgroup analysis with study design also shows nil significant moderating effect of study design (p=0.31). Subgroup analysis across geopolitical zone was not possible as only Southwest zone had sufficient study for a meta-analysis (OR 0.63, 95% CI: 0.41–0.96) as the Southeastern zone and the Northern region only had two and one studies each respectively.

#### Univariate meta-regression

Univariate meta-regression using the year of publication showed that the year of publication was significant in the association between oral hygiene status and caries with prevalence (p=0.02) Figure 5. R^2^ showed that the year of publication accounts for 100% of all the variance in study heterogeneity.

**Figure 5:**
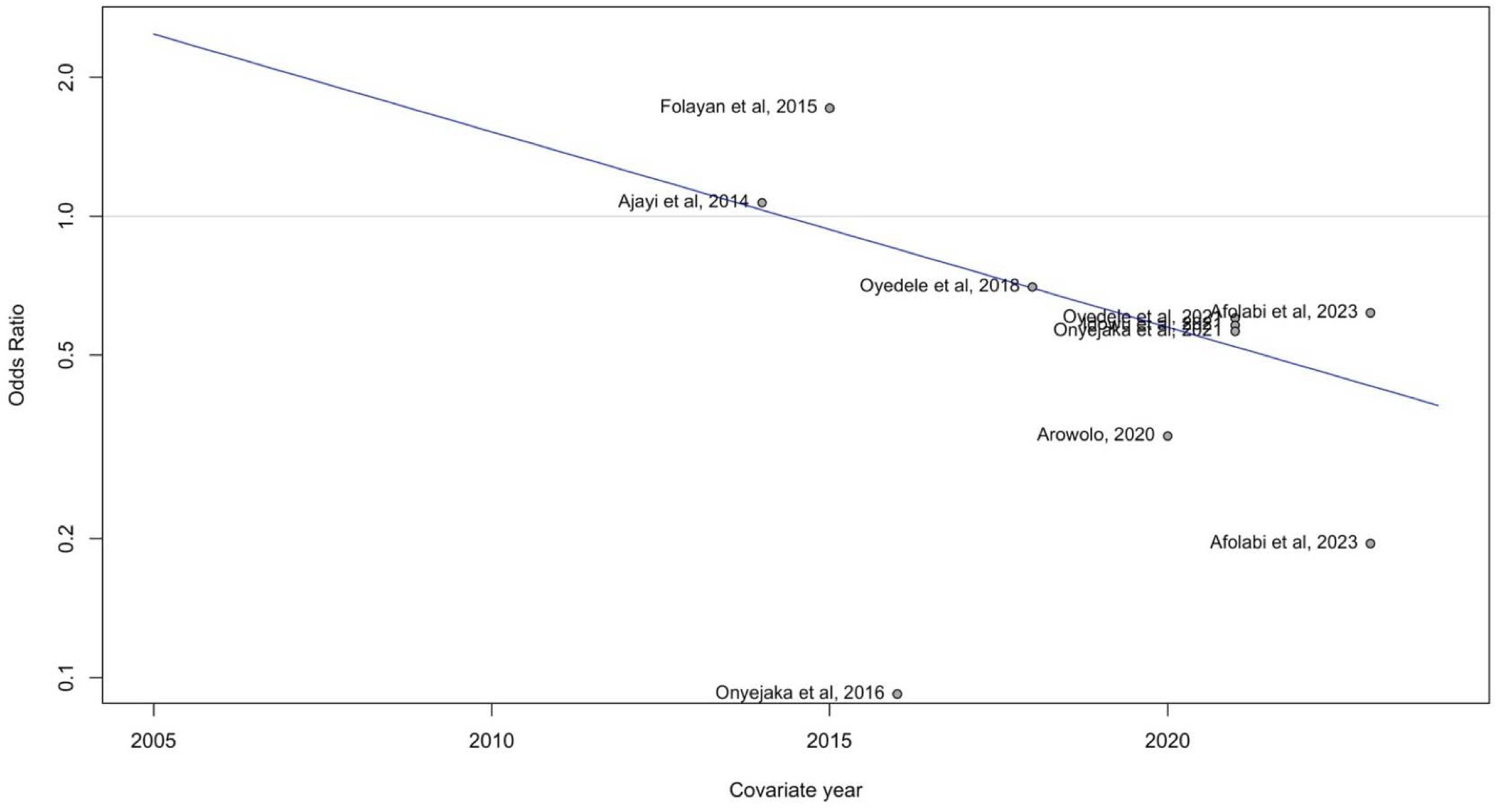
Bubble plot showing the year of publication and caries prevalence

Meta-regression also showed that the individual study sample size (p=0.31) and the mean age of participants (p=0.86) had no significant moderating effect on OH status and caries prevalence in included studies

#### Publication Bias

Because fewer than 10 included research studies were included, following the removal of the influential study, we could not evaluate the included studies’ publication bias, as the lack of statistical power from the inclusion of a few studies would cause [42].

#### Association between frequency of tooth brushing and dental caries

Analysis of 16 studies showed that brushing teeth at least twice daily was associated with a 99% reduction in the odds of dental caries compared to brushing once daily (OR 0.01, 95% CI 0.00–0.01; p < 0.001). Results were consistent across studies (I² = 0%).

**Figure 6:**
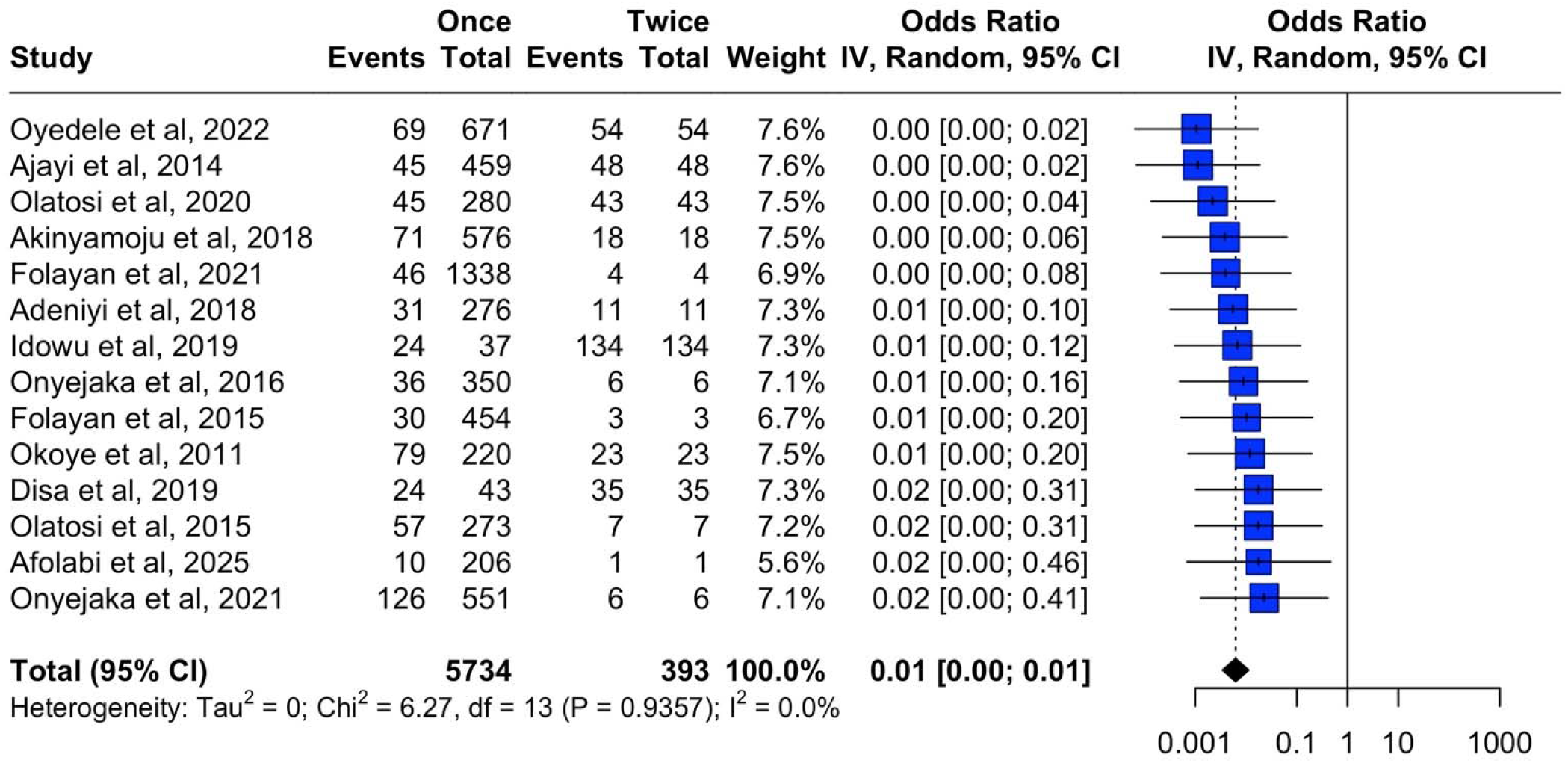
Meta-analysis showing the association of tooth brushing frequency and dental caries

### Subgroup analysis

Subgroup analysis shows no significant differences in the pooled odds ratio for tooth cleaning frequency and caries prevalence across geopolitical zones (p = 0.11), type of dentition (p = 0.24) and study design (p = 0.83)

### Publication bias

Funnel plot as shown in Figure 7 showed no funnel plot asymmetry to indicate publication bias; objectively, Egger’s test also corroborated the absence of publication bias in studies reporting dental caries prevalence and frequency of tooth brushing.

**Figure 7:**
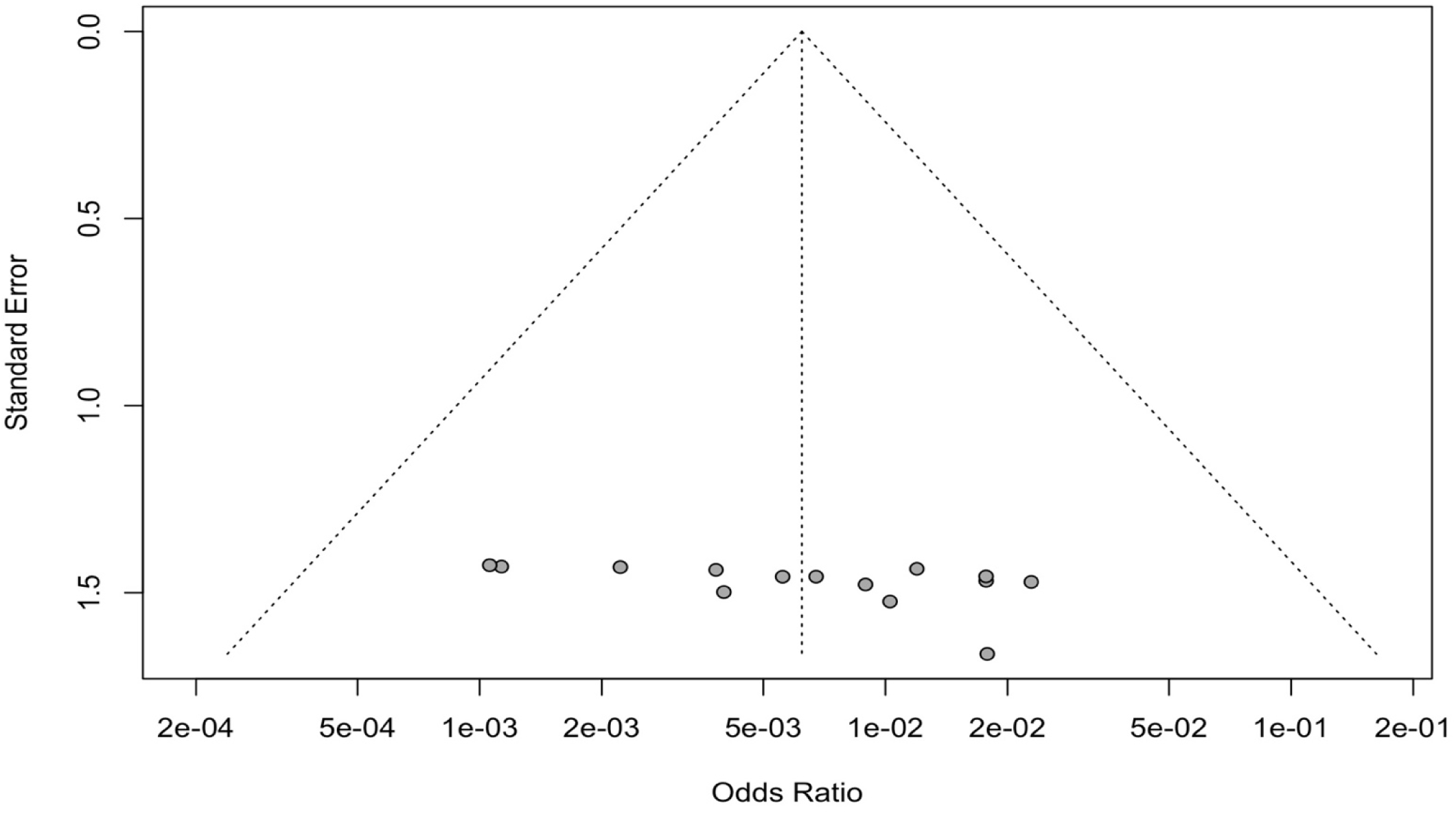
Funnel plot showing plot symmetry and lack of publication bias

### Association between tooth cleaning methods and dental caries

Analysis of eight studies found no significant association between the type of tooth cleaning device (e.g., toothbrush) and caries prevalence (OR 1.02, 95% CI 0.52–2.00; p = 0.95). Heterogeneity was high (I² = 79.5%). Sensitivity analysis indicated one influential study; its removal yielded a non-significant 19% reduction in caries odds (OR 0.81, 95% CI 0.42–1.54).

**Figure 8:**
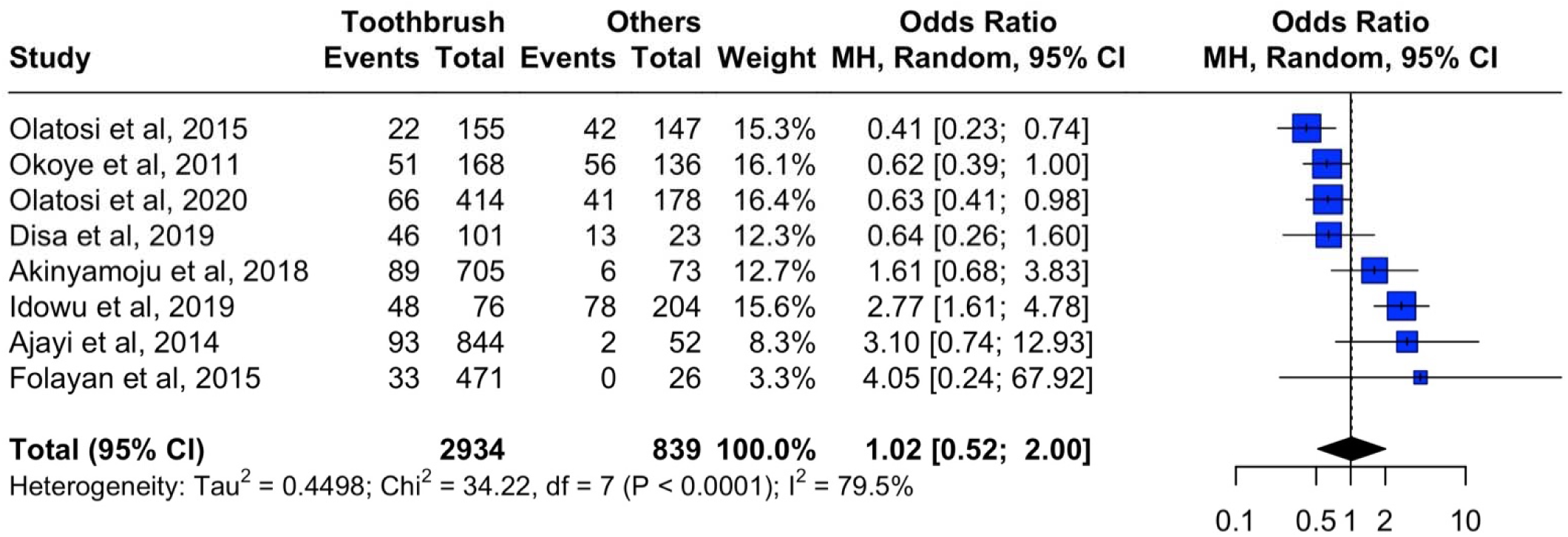
Meta-analysis showing the association of tooth brushing methods and dental caries

### Subgroup analyses

Toothbrushing was significantly associated with reduced caries prevalence in primary (42% reduction) and permanent (36% reduction) dentition, but with a 92% increase in mixed dentition (p = 0.0015). The geopolitical zones (p = 0.58) and study design (p = 0.13) exerted no significant moderating effect on the association between pooled prevalence of dental caries and tooth cleaning methods.

**Table 4:**
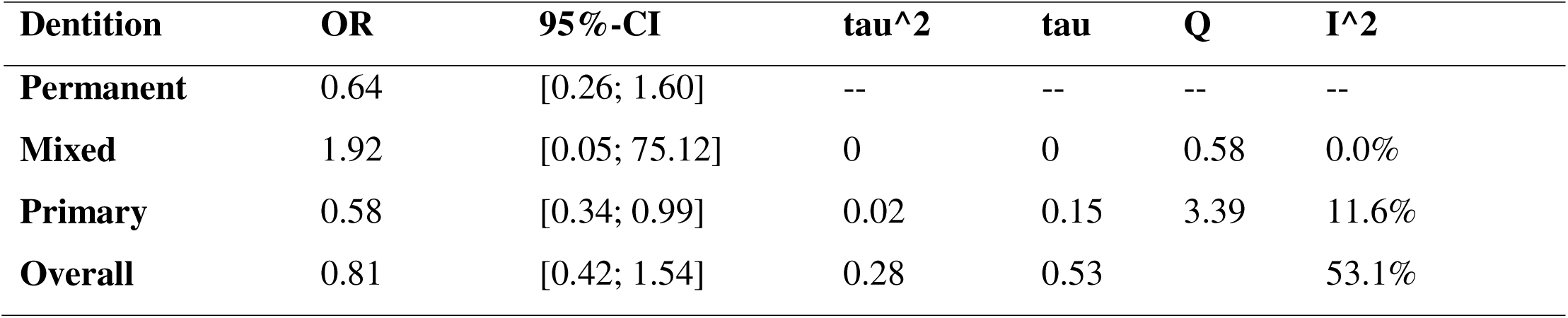
Subgroup analysis for dentition for Toothbrushing and Dental Caries: Results for subgroups (random effects model)

| Dentition | OR | 95%-CI | tau<sup>2</sup> | tau | Q | I<sup>2</sup> |
| --- | --- | --- | --- | --- | --- | --- |
| Permanent | 0.64 | [0.26; 1.60] | -- | -- | -- | -- |
| Mixed | 1.92 | [0.05; 75.12] | 0 | 0 | 0.58 | 0.0% |
| Primary | 0.58 | [0.34; 0.99] | 0.02 | 0.15 | 3.39 | 11.6% |
| Overall | 0.81 | [0.42; 1.54] | 0.28 | 0.53 |  | 53.1% |

Univariate meta-regression using sample size (p=0.043) of individual studies showed that study sample sizes were a significant moderating factor in the association between tooth cleaning methods and caries prevalence (Figure 5). R^2^ showed that sample sizes accounted for 79.8% of the study heterogeneity in the included studies.

The year of publication (p=0.30), and mean age of participants (p=0.88) had no significant moderating effect on caries prevalence and tooth cleaning methods

**Figure 9:**
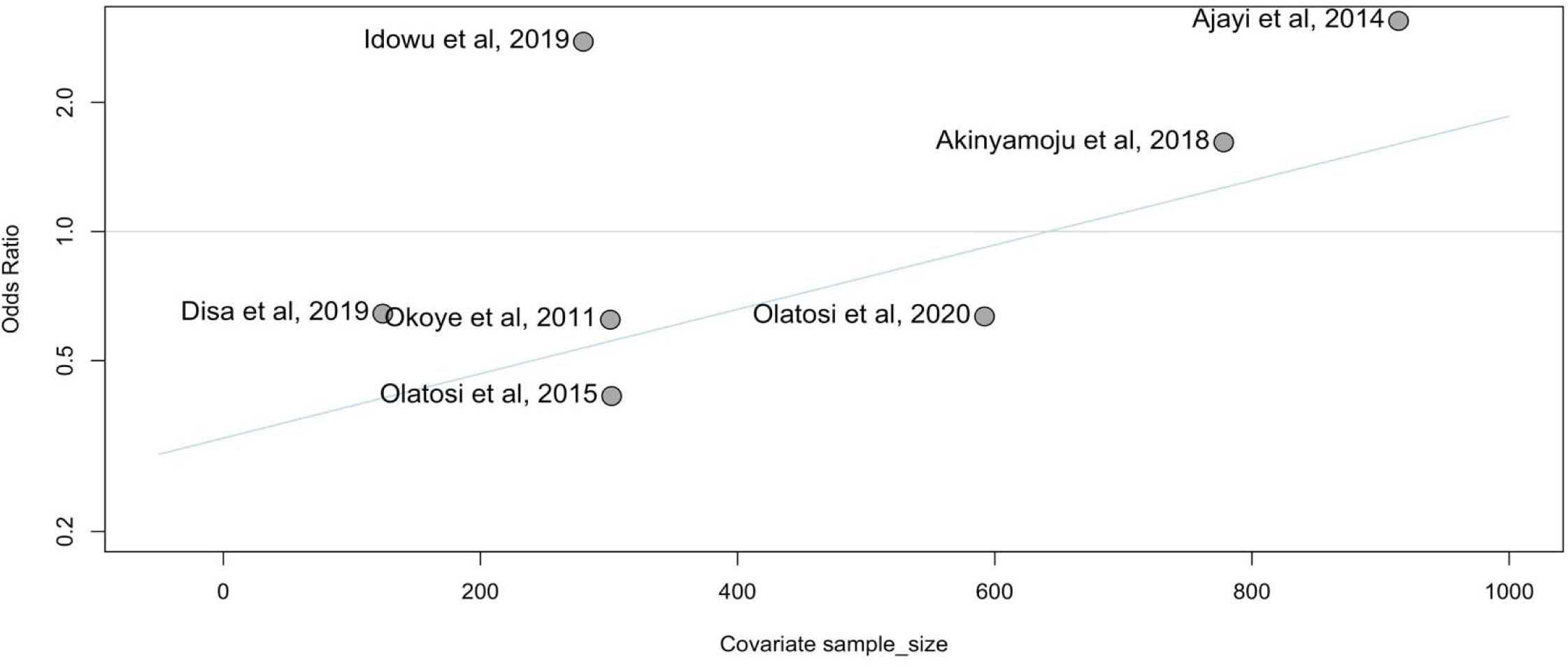
Bubble plot showing the association of tooth brushing methods and sample sizes of studies

## Discussion

This systematic review and meta-analysis present the first nationally estimated data on the associations between dental caries and oral hygiene practices. The study indicated that oral hygiene status was most commonly measured using the OHI-S, while the prevalence of caries was mainly assessed using the DMFT/dmft indices. However, the metrics for measuring oral hygiene practices were diverse, and indicators lacked uniformity across the included studies. The dental caries experience was higher when oral hygiene was poor, but this relationship was most significant in the primary dentition, and less so in the permanent or mixed dentition. In addition, toothbrushing twice daily or more was associated with a reduction in the odds of having dental caries compared to brushing once daily or less. However, there was no difference in the frequency of dental caries experience among individuals using toothbrushes and toothpaste compared to those using other tooth-cleaning devices.

A strength of the study is the methodological rigor and the transparency of the meta-analytic process, enhancing reproducibility. However, the review has several limitations that should inform the interpretation of findings and guide future research. The cross-sectional design of all included studies precludes causal inference. Unmeasured confounders such as dietary sugar intake, socioeconomic status, fluoride exposure, and access to dental care may influence the observed associations. In addition, there was a significant geographic imbalance, with most studies conducted in Southern Nigeria, particularly the Southwest. This limits the generalizability of findings to Northern Nigeria, where cultural, dietary, and socio-economic contexts may differ substantially. Future studies must prioritize this region to enable nationally representative conclusions. Despite these limitations, the study provides important information.

The first insightful finding from this systematic review and meta-analysis is the direct correlation between the prevalence of dental caries and twice-daily brushing. This contradicts simplistic biological models but becomes intelligible when considering the differential exposure to dietary sugars across socioeconomic groups, a key component of the broader caries etiology. In Nigeria, a low-middle-income country, a distinct socio-economic gradient influences oral health risk profiles: urban and higher-income populations are experiencing a nutrition transition, characterized by increased consumption of processed foods and sugary snacks and beverages [54] as seen in other transiting economies [55–59]. While these groups typically have better access to toothbrushes and toothpaste and better dental service utilisation (leading to better OHI-S scores), their high-frequency exposure to fermentable carbohydrates creates a persistent cariogenic challenge [60, 61, 62]. Here, even fair oral hygiene may be insufficient to counteract a high-sugar diet, leading to caries despite relatively better plaque scores.

On the other hand, rural and low-income populations may have more traditional, less processed diets with lower between-meal sugar intake [63]. However, they face barriers to accessing modern oral hygiene tools and preventive dental care [64]. Their oral hygiene status may be classified as poor due to plaque accumulation from less frequent or less effective cleaning methods. Yet, the lower dietary sugar exposure results in a less cariogenic oral environment, potentially leading to the observed lower caries odds. This aligns with the substrate limitation principle in cariology. Thus, the OHI-S index primarily captures plaque accumulation but not the diet’s cariogenic potential. In a setting like Nigeria with stark socioeconomic disparities, this creates an ecological fallacy: the group with better hygiene (a proxy for higher SES and urbanization) carries a higher diet-mediated risk, while the group with poorer hygiene (a proxy for lower SES and more traditional living) has a lower diet-mediated risk. This finding needs to be studied further as the trend seems to be changing among adolescents in Nigeria, wherein poorer oral hygiene is associated with higher dental caries prevalence [65].

The significant association between poor oral hygiene, dental caries, and the primary dentition also needs to be explored further, as it is currently under-explored in its full complexity because of the lifelong consequences. Primary teeth are structurally more vulnerable to rapid caries progression than permanent teeth due to key anatomical differences. The broad contact points between molars trap food and plaque allow for early caries formation. Their enamel and dentine are thinner and less mineralized, enabling faster penetration by caries [66]. While most studies focus on S. *mutans* and caries aetiology, the broader plaque microbiome and its ecological shifts (dysbiosis) in response to hygiene and diet in early childhood are an evolving area of study [67]. Further exploration must move beyond reaffirming the link and focus on developing and implementing culturally sensitive, equitable, and evidence-based strategies that disrupt this association at the individual, family, and community levels. This is an investment in a child’s immediate well-being and their long-term oral and general health trajectory.

The second insight from this systematic review and meta-analysis is the affirmation that twice-daily brushing is strongly associated with a reduced prevalence of dental caries in the study population, thereby highlighting the protective advantages of the frequency of tooth brushing in mitigating dental caries. Brushing teeth twice daily enhances plaque control by consistently disrupting the oral biofilm [68, 69]. This aligns with the conclusions made in the literature on the importance of regular brushing as a preventive measure against dental caries [68, 70]. The findings have important implications for practice, policy, and research. National oral health campaigns and school-based programs should reinforce messaging on twice-daily toothbrushing using methods identified for effectively communicating oral health [71].

The efficacy of twice-daily tooth brushing can be further enhanced by considering the optimal timing of brushing, either before or after meals, to eliminate food debris and minimize the impact of sucrose exposure through prompt cleaning after eating. [72]. In addition, using dental floss for interdental plaque removal and bacteriostatic mouthwashes can augment the effectiveness of brushing by chemically disrupting the oral biofilm. [73, 74, 75]. However, none of the studies included in this study reinforced the value of using adjuncts in reducing the caries experience, although none explored the relationship between the use of mouthwash and dental caries experience, thereby creating a knowledge gap. This gap can be bridged through future studies.

Also, although prior studies had indicated that the complementary use of fluoride-containing toothpaste with toothbrushing can further enhance the efficacy of twice-daily tooth brushing in caries control [76, 77], and local studies in the country had reinforced this [35, 78], the current study did not find such an association. This result should be interpreted cautiously and not as a contradiction of established evidence; rather, it may reflect contextual specificities or methodological limitations. The widespread use of fluoridated toothpaste in Nigeria, with over 90% of the population using it daily [67], likely created a ceiling effect in the primary studies, with such homogeneous exposure preventing the detection of a differential effect between comparison groups. These findings suggest that in a high-fluoride-toothpaste-use setting, the additional benefit of fluoride toothpaste may be obscured, and that other co-factors, such as dietary habits, may become more salient modifiers of caries risk [79].

A culturally significant finding of this meta-analysis is that toothbrushing with toothpaste was not superior to other devices in reducing the risk for caries. A population tooth cleaning tool in Nigeria is the chewing stick, drawn from plants such as *Salvadora persica* and Neem [80]. It holds deep cultural and religious significance and is traditionally believed to possess cleansing and medicinal properties [81]. The affordability and widespread availability of chewing sticks make them a sustainable alternative to toothbrushes in a resource-limited context. This study finding underscores that effective oral hygiene does not require expensive tools but rather relies on technique and consistency. A well-used chewing stick may clean more effectively than a poorly used toothbrush. This finding raises an important hypothesis that needs to be tested in future studies, challenging the longstanding assumption that modern tools are inherently superior and shifting the discussion toward a more evidence-based and culturally competent understanding of oral hygiene practices in Nigeria.

For clinicians, this finding may necessitate a shift in counselling. Rather than dismissing traditional practices, oral health professionals should engage patients with respect, asking what they use, how often, and demonstrate effective techniques. The counselling message should emphasize that the goal is efficient plaque removal, achievable with both toothbrushes and other tooth cleaning tools, while reinforcing the importance of brushing twice daily. For patients reliant on other tooth cleaning tools like chewing sticks, clinicians can encourage proper preparation and use, while complementing them with fluoride exposure through alternatives such as fluoridated mouthwash, where feasible [82]. At the community and public health level, the study evidence supports the need for campaigns that empower rather than prescribe. Messages can build on existing practices rather than attempting to replace them. This culturally sensitive approach acknowledges traditional methods while amplifying the most important public health message about frequency and thoroughness rather than the tool itself, which resonates with cultural realities and socioeconomic conditions. Further studies on using culturally appropriate oral hygiene practices are warranted to enhance our understanding of effective strategies for caries prevention in diverse populations.

In addition, the diversity observed in the array of measures employed to assess oral hygiene practices in the studies included in this systematic review may have contributed to the substantial outcome heterogeneity. It is, however, essential to study these measures to develop context-specific and relevant instruments for caries risk assessment that enable laypersons and healthcare practitioners to screen individuals at risk for caries. This gap highlights the need to develop standardized, validated tools for oral hygiene assessment in future research conducted in Nigeria.

On the other hand, we observed that the studies included in this systematic review and meta-analysis assessed dental caries using the DMFT/dmft indices. [83], a reliable and user-friendly tool for capturing dental caries experiences, with some limitations to its use [84, 85], one of which is its inability to detect enamel caries (caries in their reversible form) [86, 87]. Thus, the prevalence of caries would likely be underestimated in the studies included in this meta-analysis. Future research endeavors in Nigeria should consider transitioning to tools capable of identifying enamel caries, such as ICDAS, Cardiogram, and CAMBRIA, among others. [88].

In conclusion, this study offers a nuanced understanding of oral hygiene practices and their association with the prevalence of dental caries in Nigeria. This review synthesizes a growing body of Nigerian research to provide evidence that is both globally coherent and locally nuanced. It affirms that twice-daily brushing and good oral hygiene status are significantly associated with lower caries prevalence. It also provides data suggesting that, within the Nigerian context, the efficacy of oral hygiene may depend more on the frequency and quality of the practice than on a specific cleaning device. These findings advocate for public health strategies that promote behavioral consistency and effective techniques, while being respectful of cultural practices, to effectively reduce the burden of dental caries in Nigeria.

## Data Availability

All datasets generated and analyzed, including the study protocol, search strategy, list of included and excluded studies, data extracted, analysis plans, and quality assessment, are available in the article and upon request from the corresponding author.

## Acknowledgments

The authors thank the Nigerian Institute for Medical Research for supporting the study.

## Authors Contributions

MOF developed the idea. TG, AS, IA, MO, OA, MOF, GUE, and FTA wrote and registered the protocol on PROSPERO. UN, AOE, JL, GUE, and FTA undertook the literature searches. AA, AMA, and CO screened titles, abstracts, and full-text articles, extracted all data, and performed the initial data analysis. AA and FTA undertook a meta-analysis. MET, OCE, MOF, GUE, AA, and FTA wrote the first draft of the manuscript. All authors contributed to the writing of each manuscript draft and have approved the final submitted version.

## Ethical approval

Ethical approval was not required for this systematic review as the research was based on information retrieved from published studies.

