## Supplementary material for "Oral Hygiene Practices and Dental Caries Experience in Nigeria: A Systematic Review and Meta-Analysis": Supplemental File 1_ Search_terms_database.docx

**Supplemental File 1: Search Terms for Individual Databases**

PubMed	330

("Dental Caries"[MeSH Terms:noexp] OR ("Dental Caries"[All Fields] OR "dental decay"[All Fields] OR "oral decay"[All Fields] OR ("carie"[All Fields] OR "Dental Caries"[MeSH Terms] OR ("dental"[All Fields] AND "caries"[All Fields]) OR "Dental Caries"[All Fields] OR "caries"[All Fields]) OR ("decay"[All Fields] OR "decayed"[All Fields] OR "decaying"[All Fields] OR "decays"[All Fields]) OR "tooth decay"[All Fields] OR "dental cavities"[All Fields] OR "oral cavities"[All Fields])) AND ("Oral Hygiene"[MeSH Terms:noexp] OR ("Oral Hygiene"[All Fields] OR "oral health"[All Fields] OR "dental hygiene"[All Fields] OR "dental health"[All Fields] OR "oral practice"[All Fields] OR "dental practice"[All Fields] OR "oral habit"[All Fields] OR (("dental health services"[MeSH Terms] OR ("dental"[All Fields] AND "health"[All Fields] AND "services"[All Fields]) OR "dental health services"[All Fields] OR "dental"[All Fields] OR "dentally"[All Fields] OR "dentals"[All Fields]) AND ("habits"[MeSH Terms] OR "habits"[All Fields] OR "habit"[All Fields])))) AND ("Nigeria"[MeSH Terms:noexp] OR ("Nigeria"[MeSH Terms] OR "Nigeria"[All Fields] OR "nigeria s"[All Fields]))

Scopus	248

("Dental Caries"OR ("Dental Caries"OR "dental decay"OR "oral decay" OR ("carie" OR "Dental Caries" OR ("dental" AND "caries") OR "Dental Caries" OR "caries") OR ("decay" OR "decayed" OR "decaying" OR "decays") OR "tooth decay" OR "dental cavities" OR "oral cavities")) AND ("Oral Hygiene" OR ("Oral Hygiene" OR "oral health" OR "dental hygiene" OR "dental health" OR "oral practice" OR "dental practice" OR "oral habit" OR (("dental health services" OR ("dental" AND "health" AND "services") OR "dental health services" OR "dental" OR "dentally" OR "dentals") AND ("habits" OR "habits" OR "habit")))) AND ("Nigeria" OR ("Nigeria" OR "nigeria s"))

African Index Medicus (AIM)	10

("Dental Caries"OR ("Dental Caries"OR "dental decay"OR "oral decay" OR ("carie" OR "Dental Caries" OR ("dental" AND "caries") OR "Dental Caries" OR "caries") OR ("decay" OR "decayed" OR "decaying" OR "decays") OR "tooth decay" OR "dental cavities" OR "oral cavities")) AND ("Oral Hygiene" OR ("Oral Hygiene" OR "oral health" OR "dental hygiene" OR "dental health" OR "oral practice" OR "dental practice" OR "oral habit" OR (("dental health services" OR ("dental" AND "health" AND "services") OR "dental health services" OR "dental" OR "dentally" OR "dentals") AND ("habits" OR "habits" OR "habit")))) AND ("Nigeria" OR ("Nigeria" OR "nigeria s"))

Web of Science

("Dental Caries"OR ("Dental Caries"OR "dental decay"OR "oral decay" OR ("carie" OR "Dental Caries" OR ("dental" AND "caries") OR "Dental Caries" OR "caries") OR ("decay" OR "decayed" OR "decaying" OR "decays") OR "tooth decay" OR "dental cavities" OR "oral cavities")) AND ("Oral Hygiene" OR ("Oral Hygiene" OR "oral health" OR "dental hygiene" OR "dental health" OR "oral practice" OR "dental practice" OR "oral habit" OR (("dental health services" OR ("dental" AND "health" AND "services") OR "dental health services" OR "dental" OR "dentally" OR "dentals") AND ("habits" OR "habits" OR "habit")))) AND ("Nigeria" OR ("Nigeria" OR "nigeria s"))

African Journals Online (AJOL)	69

“dental caries AND "oral hygiene"

Google Scholar 602

"dental caries" Nigeria "oral status" OR "oral habits" "oral hygiene"

National Postgraduate College Medical College 0

"dental caries" Nigeria "oral status"

West African College of Surgeons 8

"dental caries" Nigeria "oral status"

Citation Searching

25
