## Supplementary material for "Oral Hygiene Practices and Dental Caries Experience in Nigeria: A Systematic Review and Meta-Analysis": Supplemental File 3_Assessment of Risk of Bias.docx

**Supplementary File 2: Assessment of Risk of Bias**

| **S/N** | **Author’s Name** | **Was the sample frame appropriate to address the target population?** | **Were study participants sampled in an appropriate way?** | **Was the sample size adequate?** | **Were the study subjects and the setting described in detail?** | **Was the data analysis conducted with sufficient coverage of the identified sample?** | **Were valid methods used for the identification of the condition?** | **Was the condition measured in a standard, reliable way for all participants?** | **Was there appropriate statistical analysis?** | **Was the response rate adequate, and if not, was the low response rate managed appropriately?** | **Total score** | **Grade** |
| --- | --- | --- | --- | --- | --- | --- | --- | --- | --- | --- | --- | --- |
| 1. | **Akinyamoju, 2018** | 1 | 1 | 1 | 1 | 1 | 1 | 1 | 1 | 1 | 9 | **9/9 low** |
| 2. | **Folayan, 2015** | 1 | 1 | 0 | 1 | 1 | 1 | 1 | 1 | 1 | 8 | **8/9 low** |
| 3. | **Olatosi, 2015** | 1 | 1 | 1 | 1 | 1 | 1 | 1 | 1 | 1 | 9 | **9/9 low** |
| 4. | **Onyejaka, 2021** | 0 | 1 | 0 | 1 | 1 | 1 | 1 | 1 | 1 | 7 | **7/9 low** |
| 5. | **Oyedele, 2018** | 0 | 1 | 0 | 1 | 1 | 1 | 1 | 1 | 1 | 7 | **7/9 low** |
| 6. | **Abiola, 2009** | 1 | 1 | 1 | 1 | 1 | 1 | 1 | 1 | 1 | 9 | **9/9 low** |
| 7. | **Folayan, 2021** | 1 | 1 | 1 | 1 | 1 | 1 | 1 | 1 | 1 | 9 | **9/9 low** |
| 8. | **Kolawole, 2019** | 1 | 1 | 1 | 1 | 1 | 1 | 1 | 1 | 1 | 9 | **9/9 low** |
| 9. | **Olatosi, 2015** | 1 | 1 | 1 | 1 | 1 | 1 | 1 | 1 | 1 | 9 | **9/9 low** |
| 10. | **Oyapero, 2020** | 1 | 1 | 1 | 1 | 1 | 1 | 1 | 1 | 1 | 9 | **9/9 low** |
| 11. | **Oyedele, 2022** | 1 | 1 | 1 | 1 | 1 | 1 | 1 | 1 | 1 | 9 | **9/9 low** |
| 12. | **Sowole, 2007** | 1 | 1 | 1 | 1 | 1 | 1 | 1 | 1 | 1 | 9 | **9/9 low** |
| 13. | **Okoye 2011** | 0 | 1 | 0 | 1 | 1 | 1 | 1 | 1 | 1 | 7 | **7/9 low** |
| 14. | **Ajayi 2014** | 0 | 1 | 0 | 1 | 1 | 1 | 1 | 1 | 1 | 7 | **7/9 low** |
| 15. | **Onyejaka, 2016** | 1 | 1 | 1 | 1 | 1 | 1 | 1 | 1 | 1 | 9 | **9/9 low** |
| 16. | **Idowu 2019** | 1 | 1 | 1 | 1 | 1 | 1 | 1 | 1 | 1 | 9 | **9/9 low** |
| 17. | **Disa 2019** | 0 | 0 | 0 | 1 | 1 | 1 | 1 | 1 | 1 | 6 | **6/9 Moderate** |
| 18. | **Olatosi 2020** | 1 | 1 | 1 | 1 | 1 | 1 | 1 | 1 | 1 | 9 | **9/9 low** |
| 19. | **Idowu 2021** | 1 | 1 | 1 | 1 | 1 | 1 | 1 | 1 | 1 | 9 | **9/9 low** |
| 20. | **Oyedele 2021** | 1 | 1 | 1 | 1 | 1 | 1 | 1 | 1 | 1 | 9 | **9/9 low** |
| 21. | **Folayan 2021** | 1 | 1 | 1 | 1 | 1 | 1 | 1 | 1 | 1 | 9 | **9/9 low** |
| 22. | **Arowolo, 2020** | 1 | 1 | 1 | 1 | 1 | 1 | 1 | 1 | 1 | 9 | **9/9 low** |
| 23. | **Afolabi 2023** | 1 | 1 | 1 | 1 | 1 | 1 | 1 | 1 | 1 | 9 | **9/9 low** |

**1 – 3 = High,	1 Yes**

**4 – 6 = Moderate	0 No**

**7 – 9 = Low**
